# Qatar Genome: Insights on Genomics from the Middle East

**DOI:** 10.1101/2021.09.19.21263548

**Authors:** Hamdi Mbarek, Geethanjali Devadoss Gandhi, Senthil Selvaraj, Wadha Al-Muftah, Radja Badji, Yasser Al-Sarraj, Chadi Saad, Dima Darwish, Muhammad Alvi, Tasnim Fadl, Heba Yasin, Fatima Alkuwari, Rozaimi Razali, Waleed Aamer, Fatemeh Abbaszadeh, Ikhlak Ahmed, Younes Mokrab, Karsten Suhre, Omar Albagha, Khalid Fakhro, Ramin Badii, Said I. Ismail, Asma Althani, for the Qatar Genome Program Research Consortium

## Abstract

Despite recent biomedical breakthroughs and large genomic studies growing momentum, the Middle Eastern population, home to over 400 million people, is under-represented in the human genome variation databases. Here we describe insights from phase 1 of the Qatar Genome Program which whole genome sequenced 6,045 individuals from Qatar. We identified more than 88 million variants of which 24 million are novel and 23 million are singletons. Consistent with the high consanguinity and founder effects in the region, we found that several rare deleterious variants were more common in the Qatari population while others seem to provide protection against diseases and have shaped the genetic architecture of adaptive phenotypes. Insights into the genetic structure of the Qatari population revealed five non-admixed subgroups. Based on sequence data, we also reported the heritability and genetic marker associations for 45 clinical traits. These results highlight the value of our data as a resource to advance genetic studies in the Arab and neighbouring Middle Eastern populations and will significantly boost the current efforts to improve our understanding of global patterns of human variations, human history and genetic contributions to health and diseases in diverse populations.

## Introduction

Several countries worldwide have initiated large-scale population genomics projects representing various regions from Africa, Europe, North and South America, South Asia and Australia (Gudbjartsson, Helgason, et al., 2015; Gudbjartsson, Sulem, et al., 2015; Gurdasani et al., 2019; Manolio et al., 2019; Naslavsky et al., 2020; Stark et al., 2019; Turro et al., 2020; Wu et al., 2019b). In addition to this groundbreaking work, there are also ongoing large collaborative efforts to increase diversity in human genetics, including the All of Us Research Program (Collins & Varmus, 2015), the Human Health and Heredity in Africa (H3Africa) Initiative (C. Rotimi et al., 2014) and the TOPMed Program (Taliun et al., 2021). Such studies provided valuable new insight into human disease, population structure and history of migration (Boomsma et al., 2014; Chiang et al., 2018; Francioli et al., 2014; Gurdasani et al., 2019; Okada et al., 2018; Scott et al., 2016a; Wu et al., 2019a). Despite this notable focus on diversity, there is still considerable effort needed to cover the broad diversity of world ancestries to ensure that discoveries does not conserve historical disparities and to uncover the various diseases etiologies that remain uncharacterized to date (Bentley et al., 2017; Landry et al., 2018; Mills & Rahal, 2019). The Middle-East regions are still underrepresented in the public databases (Abou Tayoun & Rehm, 2020). For instance, the latest version of gnomAD database (3.1) contains data from only 158 Middle-eastern genomes (Karczewski et al., 2020). The Qatar Genome Program (QGP) is a population genome project based in Qatar aiming to sequence the genomes of local population for the purpose of supporting genomic medicine in the country and the region. As part of phase 1, it has sequenced the whole genomes of 6,045 subjects whose specimens were collected and biobanked by the Qatar Biobank (QBB) (Al Thani et al., 2019b) Figure 1a).

**Figure 1.** Qatar Genome Program, timelines, and regional context. a) Three phases project timeline and current status. b) Qatar Geographical map. Qatar is located in the north-eastern coast of the Arabian Peninsula with an area of 11,521 km2 sharing borders with Saudi Arabia from the south and maritime borders with Bahrain, UAE, and Iran. c) The Arabian Peninsula is believed to be the first stop in human migration out of Africa, and home for the first ancient Eurasian populations, whom later spread throughout Asia and Europe.

Qatar occupies a relatively small surface area of 11,521 km^2^ on the western coast of the Arabian Gulf. Qatar shares its southern border with Saudi Arabia and a maritime border with Bahrain, UAE, and Iran (Figure 1b) and has a population of approximately 2.8 million. The country is located at a historic intersection of ancient and recent migration and admixture (Arauna et al., 2017; Hellenthal et al., 2014). Similar to other countries in the region, it is known for its unique population structure that is characterized by a high consanguinity rate and increased prevalence of rare genetic diseases (Al-Gazali et al., 2006; Anwar et al., 2014; Hunter-Zinck et al., 2010; Rodriguez-Flores et al., 2014, 2016; Scott et al., 2016a). Recent genetic studies identified indigenous Arabs as the direct descendants of the first Eurasian populations established by early migrations out of Africa (Bentley et al., 2017) (Figure 1c). Moreover, sizable proportions of the population have more recent Persian and African ancestry (Harkness & Khaled, 2014). QBB includes comprehensive phenotyping, providing excellent synergy for discovery when combined with the WGS data, that also enable accurate estimate of allele frequencies for rare and common variants, and well-defined polygenic risk scores for many disease traits. All such features of the local population potentiate discoveries, not only related to millions of people in the immediate neighboring region, but also inform genetic studies in other parts of the world.

## Materials and Methods

### Qatar Biobank subject recruitment

The Qatar Biobank (QBB) is a longitudinal population-based cohort study examining a population sample of permanent Qatari residents (Qatari nationals, other Arabs and non-Arabs) with follow up every 5 years (Al Thani et al., 2019b) To achieve a representative sample of the permanent population that resides in Qatar, the inclusion criteria of the QBB are: 1. To be Qatari nationals or resident in Qatar for at least 15 years and 2. To be 18 years or older. QBB is inclusive and language specification and tribes name or origin are not part of the inclusion criteria. The participants are recruited from the general public via either social media and the QBB website or through personal recommendations of family and friends.

The study covers extensive baseline sociodemographic data, clinical and behavioral phenotypic data, biological samples (i.e. blood, urine, saliva, DNA, RNA, viable cells and others), as well as clinical biomarkers and Omics data (i.e. genomics, transcriptomics, proteomics, metabolomics etc.) (Al Thani et al., 2019b). Currently the QBB has reached 44.7 % of the target population (60,000) and more than 2 million biological samples. For this study, data from 6,045 Qatari nationals participants were available from QBB population cohort. The percentage female was 56.74% and the mean age was 40 years (SD 12.7 years).

### Ethics Statement

All QBB participants signed an Informed Consent Form prior to their participation; QBB study protocol ethical approval was obtained from the Hamad Medical Corporation Ethics Committee in 2011 and continued with QBB Institutional Review Board (IRB) from 2017 onwards and it is renewed on an annual basis (IRB protocol number, QF-QGP-RES-PUB-002).

### Qatar Biobank sample collection

Physical and clinical measurements were collected by the QBB, in addition to biological samples (approximately 60ml of blood, 5ml of saliva, and 10ml of urine). Participants were instructed for 8 hours fasting before the visit, but due to different visit shifts samples were mostly spot specimens. Blood samples were analysed to assess 66 different biomarkers associated with disease risk factors. Haematology and blood chemistry biomarkers were analysed at Hamad General Hospital laboratories. EDTA blood samples were separated by centrifugation into plasma, buffy coat (leucocytes) and erythrocytes. All collected samples were aliquoted and stored in 3 different locations (Al Thani et al., 2019a).

### DNA isolation and Quality Control

Prior to DNA isolation, each buffy coat sample was registered into the Laboratory Information Management System (LIMS) and assigned with three identifiers: i. the aliquot code, ii.a subject-specific personal number, and iii. a sample-specific serial number. Samples were received in 2D-coded FluidX tubes (Brooks Life Sciences). Upon receiving, samples were scanned on a 2D FluidX Perception Barcode Reader to check for consistency against the sample submission form. The buffy coat samples were processed for DNA isolation using the automated QIASymphony SP instrument according to Qiagen MIDI kit protocol’s recommendations. The assessment of DNA quantity and quality was carried out using NanoDrop 8000 (Thermofisher, Waltham, MA, USA), FlexStation 3 (Molecular Devices, Sunnyvale, CA, USA) and LabChip GX (Perkin Elmer, Waltham, MA, USA). The absorbance at 260 and 280 nm wavelength was measured on Nanodrop 8000 and used to check DNA purity. A fluorescence-based quantification was performed on FlexStation 3 using Quant-iT PicoGreen dsDNA Assay (Thermofisher). Briefly, an aqueous working solution of the Quant-iT PicoGreen reagent was prepared on the day of the quantification experiment by making a 200-fold dilution of the concentrated DMSO solution in TE. TE buffer was also used for diluting DNA samples and in the assay itself. Sample measurement on FlexStation 3 was performed following the manufacturer’s recommendations. DNA integrity was checked on LabChip GX. The Gel-Dye solution, DNA samples and DNA ladder were prepared according to the manufacturer’s instructions; the run data was compared to the electropherogram of a typical high molecular weight ladder and assessed for quality. A genomic DNA (gDNA) quality score (GQS) was calculated for each sample. The GQS is derived from the size distribution of the gDNA and it represents the degree of degradation of a given sample, with a score of 5 corresponding to intact gDNA and a score of 0 corresponding to a highly degraded gDNA. Figure S1 shows the GQS distribution across 50 samples assessed from phase I. The distribution shows GQS>3.5.

### Whole genome sequencing

Library construction and sequencing was performed at the Sidra Clinical Genomics Laboratory Sequencing Facility. After extraction of genomic DNA, sample integrity was controlled using the Genomic DNA assay on the Perkin Elmer Caliper Labchip GXII. Concentration was measured using Invitrogen Quant-iT dsDNA Assay on the FlexStation 3. Around 150ng of DNA were used for library construction with the Illumina TruSeq DNA Nano kit. Each library was indexed using the Illumina TruSeq Single Indexes. Library quality and concentration was assessed using the DNA 1k assay on a Perkin Elmer GX2. Libraries were quantified using the KAPA HiFi Library quantification kit on a Roche LightCycler 480. Flow cells were loaded at 1 sample per lane and cluster generation was performed on a cBot 1.0 or 2.0 using the HiSeq X Ten Reagent Kit v2.5. Flow cells were loaded at a cluster density between 1255 and 1412 K/mm2 and sequenced on an Illumina Hiseq X instrument to a minimum average coverage of 30x.

### Sequencing data processing methods

The Sidra Bioinformatics Core (SBC) developed a pipeline to perform the NGS analysis for QGP and other internal projects (Figure S2). The core also developed a framework to automate the processing of the samples. Data is received from the clinical genomic lab (CGL) in Fastq^1^ format. Quality control of Fastq files is performed using FastQC(v0.11.2)^2^, to calculate quality metrics and ensure that raw reads have good quality. Reads are then trimmed and aligned to hs37d5^3^ reference genome using bwa.kit (v0.7.12)^4^ and a bam^5^ file is generated. Quality control on mapped reads (BAM files), to evaluate the coverage of each sample, is performed using Picard (v1.117) [CollectWgsMetrics]^6^. The variant calling is performed following GATK 3.4 best practices^7^: Indel realignment and base recalibration (BQSR) is performed on the initial bam then HaplotypeCaller run on each sample to generate an intermediate genomic gVCF (gVCF). Joint Genotyping is performed using all generated gVCF files at once. We first run GenomicsDB^8^ to combine the different samples by regions, then on each region, we run GenotypeGVCFs, apply SNP/Indel recalibration (VQSR) and then merge all regions. Annotation is performed using SnpEff/SnpSift^9 (^v4.3t). The following databases are used within SnpEff/SnpSift for the annotation of the multi-samples VCF file:

- dbSNP build 151
- ClinVar 2019-02-11
- dbNSFP^10^ v2.9
- GWAS catalog^11^
- msigDBdb^12^ v5.0

All variants are kept within the VCF file. Copy Number Variation analysis was performed using Canvas^13^ (v1.11.0) and structural variant analysis was performed using Manta^14^ (v0.29.6) and Delly^15^ (v0.7.8). Both analyses use bam file as input and were performed at the single sample level. Additionally, QGP VCF file was decomposed for multi allelic position and then normalize using vt^16^ (v0.5). QGP VCF file was split chromosome wise and this per chromosome VCF file was provided for further analysis as well. All pipeline references are in the supplemental data.

To identify disease-causing variants in HGMD, ClinVar and OMIM, we used VCF file annotated with phenotype/disease information from these databases. To achieve that, we applied successive filtering on the variant list using different criteria (selecting only those located in known HGMD/OMIM gene, variants with MAF <1% in all databases, except QGP, and the variant should be within or affecting the coding region; missense, nonsense, frameshift, and splice-site variants). Among the final list, we selected those that have been previously reported and flagged as disease-causing “DM/DM?” in HGMD or “Pathogenic/Likely_pathogenic” in ClinVar.

### Data Quality Control

QGP phase I study included 6,218 samples. We applicate downstream quality control on the multi-sample VCF using the PLINK v2.0 tool (Chang et al., 2015). After quality control, 8 samples were removed for excess heterozygosity, 1 for low-call rates (less than 95%), 65 for gender mismatch, 87 for population outliers (individuals with more than four standard deviation (±4 SD) away from the mean of the first two multidimensional scaling component), and 10 for identical matching. After these exclusions (N= 171), a final set of 6,045 subjects was obtained for which whole-genome sequencing was performed at a median depth of 32X (Thareja et al. (in press).

### Statistical analyses

We compared the allele counts of QGP samples to allele counts present in gnomAD exome samples for HGMD DM variants. A Fisher’s exact test was used to calculate variations that were significantly overrepresented in the QGP samples (due to founder effect) and corrected for multiple testing using the Bonferroni method.

### Hail genomic processing tool

Data preprocessing and analysis was performed using Hail 0.2. allele count, allele number, allele frequency, homozygous count calculation for each subpopulation was performed simultaneously using python scripts written using hail framework. Quality analysis for variant calls and individual sample were performed using variant_qc and sample_qc functions respectively. Sample level statistics for each sample was generated using the Hail.

### QGP variant browser

QGP variant browser provides a mechanism for the researchers to be able to search, filter and browse the QGP genomic variants data. This web-based browser supports fast database query response time for searching through more than 88 million records with search and filter functionality on the QGP gene variants and its attributes (e.g. allele frequency, homozygosity etc.).

## Results

### Genetic variability of the Qatari population

We have identified a total of 88,191,239 variants, which includes 74,991,446 SNVs (74,040,559 bi-allelic SNVs) with 939,405 multi-allelic sites and 13,199,792 INDELS (8,389,562 bi-allelic INDELS) with 2,018,185 multi-allelic sites/microsatellites (Figure 2a-c and Figure S3). Importantly, twenty-eight percent (28%) of the total variants (24,620,313) were novel and not previously reported in dbSNP build 151 or other population databases (gnomAD, 1000 Genomes, and Greater Middle East (GME)) (Figure 2b; Figure S4a-b and 5). Each individual genome presented a median of 3.4 million SNVs and 63,755 novel variants. We estimated the transition to transversion (ti/tv) ratio of 2.05 and heterozygotes to non-ref homozygote (Het/Hom) ratio of 1.85, which is consistent with previous WGS studies (Auton et al., 2015). We found 23 million variants present as singletons which are less when compared to the number of variants falling under the minor allele frequency (MAF) spectrum of <0.1% (2-12 alleles) which should be around 34 million variants (Figure2c and Table S1). While considering the novel variants, singletons (45%) being slightly higher than the variants that fall in the category of 2-12 alleles (42%) and only 13% of the novel variants exceed the MAF > 0.1%. Half of the singletons present in QGP were already reported in dbSNP and, each individual carried a median of 1,336 singletons (Figure 2d and Figure S6).

**Figure 2.** Variants distribution and allele frequency spectrum of QGP data. a) Number of SNVs and INDELS present within the QGP data. b) Known and novel variants distribution of QGP data. c) QGP variants classification based on minor allele frequency (MAF). d) Proportion of known and novel singletons within the QGP data. e) Classification of DM variants based on pattern of inheritance. Inheritance patterns of genes were derived from OMIM database. f) Distribution of DM variants among individuals in QGP sub clusters. g) QGP variants classified as both DM and pathogenic/likely pathogenic.

To evaluate the impact and scale of disease-causing variants in our population, we annotated the variant list with disease/phenotype information from HGMD, ClinVar and OMIM databases. In total, we found 4,254 disease-causing mutations (DM), which includes 3,970 SNVs and 284 INDELS (Figure S7a). These variants are located across 1,672 genes that are linked to phenotypes with different modes of inheritance (678 follow autosomal recessive (AR); 315 autosomal dominant (AD); 526 both AR and AD; and 50 X-linked inheritance) (Figure 2e). The vast majority (97%) of these DM variants are rare with MAF <1%, and among them 30% observed as singletons (Figure S7b). Each individual in the QGP dataset carries a median of 21 DM variants (range of 8-37) (Figure 2f and Figure S7c), slightly less than what have been previously reported (25 DMs/individual in the UK10k (Xue et al., 2012) and 29 DMs/individual in the Uganda genome studies (Gurdasani et al., 2019)). Each individual also carries in the homozygous state a median of 5 DM variants (range of 1-11) compared to 3 homozygous DMs/individual in the Uganda genome and 3–24 homozygous variants in the 1000 Genome project (Auton et al., 2015; Gurdasani et al., 2019). Our data shows that approximately 900 protein-coding genes have at least 1 DM mutation and 26 genes present 15 or more DM mutations (Figure S7d). When QGP data is classified according to ClinVar information (version February 11^th^ 2019), we found that 1,449 variants are classified as “pathogenic” or “likely pathogenic” (Figure S7e). Further classification considering both HGMD and ClinVar, revealed that 1,011 variants were marked as DM and “pathogenic or likely pathogenic” (Figure 2g), with 160 variants unique to the Qatari population. Interestingly, only a subset of 14 variants, among the 1,011 variants, are shared between the QGP samples and data from Greater Middle East (GME) Variome Project (Scott et al., 2016b) (Table 1). There are also 36 variants which confer protection against several diseases including malaria, obesity, and heart disease (Table S2).

**Table 1.** Pathogenic variants unique to the Middle East region. Pathogenic variants exclusively reported in QGP and GME (Greater Middle East) variome project. QGP_MAF - Minor allele frequency in QGP data; GME_MAF – Minor allele frequency in GME variome project.

| Chrom | Pos | ID | QGP_MAF | GME_MAF | GENE | HGVS_C | HGVS_P | Annotation | HGMD | CLINVAR | Disease Phenotype |
| --- | --- | --- | --- | --- | --- | --- | --- | --- | --- | --- | --- |
| 2 | 47604159 | rs606231204 | 0.00223251 | 0.000503525 | EPCAM | c.583dupC | p.Gln195fs | frameshift variant | DM | Pathogenic | Congenital Tufting Enteropathy (CTE) |
| 2 | 73676380 | rs746640196 | 0.000165399 | 0.000503525 | ALMS1 | c.2723C>G | p.Ser908* | stop gained | DM | Likely pathogenic | Alstrom syndrome |
| 2 | 172305304 | rs797045038 | 0.00727634 | 0.001007049 | DCAF17 | c.436delC | p.Ala147fs | frameshift variant | DM | Pathogenic | Woodhouse-Sakati syndrome |
| 3 | 113119479 | rs866096259 | 0.00330961 | 0.000503525 | WDR52 | c.1387G>T | p.Glu463* | stop gained | DM | Pathogenic | Spermatogenic failure |
| 4 | 108866582 | rs397514513 | 0.00223251 | 0.001510574 | CYP2U1 | c.947A>T | p.Asp316Val | missense variant | DM | Pathogenic | Spastic paraplegia |
| 4 | 119736287 | rs730882211 | 0.00148834 | 0.001007049 | SEC24D | c.700G>C | p.Gly234Arg | missense variant | DM | Likely pathogenic | Intellectual disability/Seizures |
| 6 | 135776888 | rs121434350 | 0.000165399 | 0.001009082 | AHI1 | c.1328T>A | p.Val443Asp | missense variant | DM | Pathogenic/Likely pathogenic | Joubert syndrome |
| 8 | 145741257 | . | 8.27E-05 | 0.000503525 | RECQL4 | c.1149G>A | p.Trp383* | stop gained | DM | Pathogenic | Rothmund-Thomson syndrome |
| 9 | 111899809 | rs878853280 | 0.000413497 | 0.001007049 | FRRS1L | c.961C>T | p.Gln321* | stop gained | DM | Pathogenic | Epileptic encephalopathy |
| 15 | 65295453 | rs863224897 | 8.27E-05 | 0.000503525 | MTFMT | c.1116delT | p.Pro373fs | frameshift variant | DM | Likely pathogenic | Moyamoya disease |
| 16 | 77369781 | rs148319220 | 8.27E-05 | 0.001007049 | ADAMTS18 | c.1731C>G | p.Cys577Trp | missense variant | DM | Pathogenic | Microcornea, myopic chorioretinal atrophy, and telecanthus(MMCAT) |
| 19 | 11304215 | . | 0.00256325 | 0.000504032 | KANK2 | c.541A>G | p.Ser181Gly | missense variant | DM | Pathogenic | Nephrotic syndrome,16 |
| 21 | 47805894 | rs387906928 | 8.27E-05 | 0.000503525 | PCNT | c.3460G>T | p.Glu1154* | stop gained | DM | Pathogenic | Microcephalic osteodysplastic primordial dwarfism type 2 (MOPD2) |
| 22 | 27012112 | rs1064793935 | 0.000248057 | 0.000504541 | CRYBB1 | c.171delG | p.Asn58fs | frameshift variant | DM | Pathogenic | Cataract |

We found some rare pathogenic variants present in Qatari population with high minor allele frequencies due to the founder effect. Some of the examples include variant in the MPL gene [MIM: 604498] (rs750046020), previously associated with thrombocytosis, occurs at a MAF of 0.009, and similarly, variants in the genes *CBS [MIM:*236200] (rs398123151) and *KRT5* [MIM: 148040](rs267607448) associated with homocystinuria and Epidermolysis Bullosa, respectively, are observed at a MAF of 0.007”.

### Genetic Ancestry and Diversity of the Qatari population

To capture the genetic diversity of the Qatari population and understand its relationship with the world’s populations in both modern and ancient times, we identified five major ancestries: General Arabs (QGP_GAR, 38%), Peninsular Arabs (QGP_PAR, 17%), Arabs of Western Eurasia and Persia (QGP_WEP, 22%), South Asians (QGP_SAS, 1%), Africans (QGP_AFR, 3%) and Admixed (QGP_ADM, 19%) (Razali et al., in press) (Figure S8). We also characterized a group of Peninsular Arabs forming a unique cluster within known descendants originating from the historical homeland of ancient Arab tribes. Analysis of Mitochondrial DNA (mtDNA) and Chromosome Y (Chr Y) in the dataset has enriched the poorly characterized landscape of haplogroups in Arab and Middle East populations in general. Notably, J1a2b a Chr Y haplogroup seen previously in Yemen, has been observed in 1,419 males, which is the largest set of individuals ever sequenced within this haplogroup. We discovered 103 novel Y-Chr SNPs in these individuals, which aided the expansion of this haplogroup to 29 novel sub-haplogroups. Using this unique dataset, we built a panel for genotype imputation for Arabs and Middle Eastern ancestries which shows an improved imputation score for rare and common allele frequencies variants (Razali et al., in press).

We next characterized the spectrum of genetic variability based on the fine-scale population structure observed in the Qatari population. This analysis highlighted that 70% of the novel variants are cluster-specific, 5% are found in all sub-clusters, and the remaining 25% are shared between one or more sub-clusters (Figure S9a). Similarly, we found that about half (2,139) of the DM variants are cluster-specific and only 68 out of 4,254 DM variants were present in all sub-clusters (Figure S9b). Furthermore, individuals in the QGP_AFR sub-cluster have the highest heterozygotes to non-ref homozygote (Het/Hom) ratio, whereas the ratio was found to be lowest for the QGP_PAR cluster. This reflects the high homozygosity and high consanguinity present within the individuals of this cluster (Figure S9c). Similarly, the median number of singletons is lower for PAR cluster compared to other sub-clusters reflects the closely related individual present in this cluster (Table 2).

**Table 2.** Median number of variant sites per genome. Novel SNV and INDELS: Variants, which are not reported in dbSNP or gnomAD or 1000G project. GERP (Genomic Evolutionary Rate Profiling) score: Scores >3 represent highly conserved positions. ADM- Admixed, AFR- Africans, GAR- general Arabs, PAR- Peninsular Arabs, SAS- South Asians, WEP- Arabs of Western Eurasia and Persia.

| Annotation | QGP<br>(n=6,045,<br>Depth = 32.4x) | ADM<br>(n=1,180,<br>Depth =32.2x) | AFR (n=92,<br>Depth=31.9x) | GAR (n=2,311,<br>Depth =32.2x) | PAR (n=1,052,<br>Depth=32.2x) | SAS (n=38,<br>Depth =31.9x) | WEP (n=1,372,<br>Depth=32.2x) |
| --- | --- | --- | --- | --- | --- | --- | --- |
| SNV | 3,467,270 | 3,596,354 | 3,967,082 | 3,466,051 | 3,391,850 | 3,492,506 | 3,458,604 |
| INDELS | 1,107,288 | 1,128,043 | 1,207,016 | 1,105,836 | 1,094,173 | 1,113,900 | 1,101,075 |
| Singletons | 1,336 | 9,242 | 17,606 | 2,484 | 408 | 20,056 | 3,193 |
| Novel SNV | 18,453 | 21,311 | 25,993 | 16,419 | 12,022 | 23,814 | 20,788 |
| Novel INDELS | 45,756 | 46,263 | 48,406 | 45,752 | 46,061 | 46,195 | 45,107 |
| Synonymous | 10,657 | 11,094 | 12,285 | 10,643 | 10,372 | 10,768 | 10,635 |
| Missense | 10,681 | 11,241 | 12,464 | 10,921 | 10,684 | 10,997 | 10,895 |
| Intron | 1,617,713 | 1,659,957 | 1,833,502 | 1,618,061 | 1,586,985 | 1,632,466 | 1,613,603 |
| Intergenic | 1,760,161 | 1,806,271 | 1,989,241 | 1,759,321 | 1,726,938 | 1,774,147 | 1,756,828 |
| Conserved: GERP>3 | 3,751 | 3,859 | 4,233 | 3,755 | 3,667 | 3,788 | 3,738 |

Furthermore, runs of homozygosity (ROH) analysis of the QGP done by Razali et al 2021 (Razali et al., in Press), identified per population ROH boundary for short, medium and long ROH. We observed that Peninsular Arabs (PAR) have the lowest median for short ROH after African-based populations. In addition, PAR has the highest median for long ROH, indicating recent consanguinity events. When we analyzed the relationship between genes and the ROH regions, we observed that there are more OMIM genes in ROH regions compared to non-OMIM genes regardless of the ROH classes. PAR was shown to have significantly more OMIM genes compared to the other QGP and 1KG populations.

### Burden of Pathogenic Variation

We then focused on the burden of pathogenic variants of recessively inherited disorders in the Qatari population. We found the most common recessive alleles are those linked to structural deformities and developmental disorders, consistent with the fact that such recessive traits prevail in societies where endogamy and consanguinity is practiced (Table S3). However, some of these identified alleles are too common to be classified as pathogenic variants (rs201818754, rs373804633, rs199768740, and rs80358230) as their frequencies in PAR subpopulation exceeding 4%, far more than the associated disease prevalence.

A notable example of an autosomal recessive disorder is Woodhouse-Sakati syndrome [WSS (MIM:241080)], a disease characterized by hypogonadism and hair thinning that often progresses to alopecia totalis. Of the less than 100 individuals reported globally with the disease, 30 are from Middle Eastern families (Bohlega & Alkuraya, 1993). WSS is caused by biallelic pathogenic variants in the [DCAF17 (MIM: 612515)] (previously known as C2orf37) gene. We identified NM_025000.4 (DCAF17): c.436delC (p.Ala147fs) as the sole pathogenic variant of this gene in 88 individuals, in heterozygous state (MAF = 0.007) (Supplementary data). Although all heterozygous individuals were found to be clinically asymptomatic, the alternate allele in these individuals is associated with the decreased levels of Insulin (Pvalue = 2.9E-02; β = -0.225; Figure S10) which could explain diabetes mellitus being one of the characteristic clinical phenotypes in WSS. We also found that c.436delC is enriched (fisher exact test P=7.57E-34; OR=18.45) in one of the founder populations, QGP_PAR subcluster, this is consistent with a previous report that identified DCAF17:c.436delC (rs797045038) as a founder variant in the Qatari population (Ben-Omran et al., 2011). This variant has also been reported in the Kingdom of Saudi Arabia (Alazami et al., 2008), which has a large number of tribes sharing common and similar carrier frequency with Qatar’s native population. Hamad Medical Corporation (HMC) is hosting the national molecular diagnostic laboratories of Qatar, and has identified to date 34 WSS patients and 64 heterozygous carriers. Data from both QGP and HMC laboratories indicates that the carrier frequency for WSS in the Qatari population is approx. 1 in 42 individuals (2.5%) with MAF of 1.25%, which is the highest reported in the world.

Remarkably, the carrier frequency of c.436delC (p.Ala147fs) is 7x higher in Qatar than in the same tribe living in neighboring Saudi Arabia and has not yet been reported in population frequency databases, such as gnomAD and 1000 genomes or the 100K Genomes Project that includes patients with rare genetic diseases (Turnbull et al., 2018).

### Insights into the genetics of quantitative traits

To gain insights into the genetic architecture of health and disease-related quantitative traits, we performed the first genome wide association studies of a list of 45 quantitative traits in 6,045 individuals from the Qatari population (Thareja et al., 2021). Several important findings of this comprehensive study include replication of multiple associations reported in Caucasian and Asian GWASs; uncovering differences in allele frequencies and LD patterns for replicated loci; and discovery of novel genetic associations mostly with variants common in the QGP but rare in other populations. These findings argue for larger GWAS studies from the region to accurately derive polygenic risk scores optimized for Middle Eastern populations for improved application in precision medicine.

## Discussion

Here we characterized a broad spectrum of genetic variation in the Qatari population, in total over 88 million variants (1.86 % of novel variants per individual genome and 24.6 M novel variants in the whole dataset). This large-scale study allowed us to identify five non-admixed subgroups in QGP (n=6,045) compared to three in the previous study Fakhro et al. 2016 (n=1,005) (Fakhro et al., 2016). We found a larger number of DM variants carried per individual which could be explained by incomplete penetrance, or the individual might carry them in a heterozygous state (Francioli et al., 2014; Xue et al., 2012). We described the distribution of genetic variation across the sub-clusters and found the majority of the novel variants to be cluster-specific. This data support records of high consanguinity and founder effect but also identify a previously unstudied component of the Middle Eastern population. Based on these sequencing results of 6,045 individuals we have recently reported a total of 60 pathogenic and likely pathogenic in 25 ACMG genes in 141 unique individuals (Elfatih et al., 2021) and several other efforts are currently under way to build the catalogues of predicted loss-of-function variants and mendelian disorders mutations and to characterize the pharmacogenomic and the cancer landscapes of the Qatari population. Furthermore, using a combination of whole genomes and exome sequence data and clinical reports, we developed a microarray with Qatari-specific pathogenic variants that could be used to rapidly, accurately and at low cost, screen the Qatari population for pathogenic variants of newborns, premarital couples and patients presenting to the clinic (Rodriguez-Flores, in press).

Previous genetic studies in the Middle East region have assessed the genomic variations linked to health and diseases mostly limited to whole exome sequencing on relatively small sample size (AlSafar et al., 2019; Fattahi et al., 2019; John et al., 2018; Monies et al., 2019; Scott et al., 2016b). Our QGP data have a key advantage over these studies since we are performing large-scale population sequencing using a whole genome approach. Although our work provided various insights into the genomic of the Middle East, we should address one limitation of our approach is that we are including only Qatari nationals in the first phase. To overcome this limitation, we are including long term residents in our next freezes.

In conclusion, this first phase of the QGP constitutes the largest comprehensive analysis of whole genomes representative of tens of millions of Arabian Peninsula and Middle East inhabitants. Such genetic information is largely lacking in global databases (Easteal et al., 2020). Our next phases will focus on specific diseases relevant to the Qatari population’s health burden - e.g. cancer, diabetes and rare diseases - while accelerating the ability to use the genome sequencing data into clinical implementation. We anticipate our data will represent a valuable resource to advance genetic studies in the Arab and neighbouring Middle Eastern populations and will significantly boost the current efforts to improve our understanding of global patterns of human variations, human history and genetic contributions to health and diseases in diverse populations (C. N. Rotimi & Adeyemo, 2021).

## Data Availability

The informed consent given by the study participants does not cover posting of participant level
phenotype and genotype data of Qatar Biobank/Qatar Genome Project in public databases. However, access to QBB/QGP data can be obtained through an established ISO-certified process by submitting a project request at https://www.qatarbiobank.org.qa/research/how-to-apply which is subject to approval by the QBB IRB committee.

https://www.qatarbiobank.org.qa/research/how-apply

## Supplemental Information

Supplemental information includes ten figures (S1–S10), three tables (S1–S3), information about *DCAF17* founder mutation, and references for the pipelines.

## Declaration of Interests

The authors declare no competing interests.

### The Qatar Genome Program Research Consortium

**Qatar Genome Project Management:** Said I. Ismail^1^, Wadha Al-Muftah^1^, Radja Badji^1^, Hamdi Mbarek^1^, Dima Darwish^1^, Tasnim Fadl^1^, Heba Yasin^1^, Maryem Ennaifar^1^, Rania Abdellatif^1^, Fatima Alkuwari^1^, Muhammad Alvi^1^, Yasser Al-Sarraj^1^, Chadi Saad^1^, Asmaa Althani^1,16^

**Biobank and Sample Preparation:** Eleni Fethnou^2^, Fatima Qafoud^2^, Eiman Alkhayat^2^, Nahla Afifi^2^

**Sequencing and Genotyping group:** Sara Tomei^3^, Wei Liu^3^, Stephan Lorenz^3^

**Applied Bioinformatics Core:** Najeeb Syed^4^, Hakeem Almabrazi^4^, Fazulur Rehaman Vempalli^4^, Ramzi Temanni^4^

**Data Management, Advanced Applications and Computing Infrastructure groups:** Tariq Abu Saqri^5^, Mohammedhusen Khatib^5^, Mehshad Hamza^5^, Tariq Abu Zaid^5^, Ahmed El Khouly^5^, Tushar Pathare^5^, Shafeeq Poolat^5^, Shafqat Baig5, Anwar Haque5, Mohamed Jama5, Rashid Al-Ali^5^

**Genetic Variability group:** Geethanjali Devadoss Gandhi^6,8^, Senthil Selvaraj^6^, Najeeb Syed^4^, Xavier Estivill^6^, Hamdi Mbarek^1^

**Population Structure and Genome Reference group:** Rozaimi Mohamad Razali^6^, Juan Rodriguez-Flores^17^, Elbay Aliyev^6^, Haroon Naeem^6^, Waleed Aamer^6^, Andrew Clark^18^, Khalid Fakhro^6^, Younes Mokrab^6^

**GWAS group:** Gaurav Thareja^7^*, Yasser Al-Sarraj*^1,8^, Aziz Belkadi^7^, Maryam Almotawa^9^, Karsten Suhre^7+^, Omar Albagha^+8,15^ (^*^equally contributed, ^+^ jointly supervised)

**Mendelian Disorders group:** Waleed Aamer^6^, Alya Al-Kurbi^6^, Aljazi Al-Maraghi^6^, Geethanjali Devadoss Gandhi^6,8^, Najeeb Syed^4^, Khalid Fakhro^6^

**Loss of Function group:** Fatemeh Abbaszadeh^10^*, Ikhlak Ahmed^5^*, Najeeb Syed^4^, Mohammad Abuhaliqa^10^, Rashid Al Ali^5^, Khalid Fakhro^6^, Zafar Nawaz^10^, Ajayeb Al Nabet Al Marri^10^, Xavier Estivill^6^, Puthen V. Jithesh^8^, Ramin Badii^10^ (^*^equally contributed)

**Consortium Lead Principal Investigators (in alphabetical order):** Omar Albagha^8,15^, Souhaila Al-Khodor^11^, Mashael Alshafai^12^, Ramin Badii^10^, Lotfi Chouchane^13^, Xavier Estivill^6^, Khalid Fakhro^6^, Hamdi Mbarek^1^, Younes Mokrab^6^, Puthen V. Jithesh^8^, Karsten Suhre^7^, Zohreh Tatari^14^

### Affiliations

1. Qatar Genome Program, Qatar Foundation Research, Development and Innovation, Qatar Foundation, Doha, Qatar.
2. Qatar Biobank for Medical Research, Qatar Foundation, Building 317, Hamad Medical City, Doha, Qatar.
3. Sidra Medicine, Integrated Genomics Services, Out-Patient Clinic, Doha, Qatar.
4. Sidra Medicine, Applied Bioinformatics Core - Integrated Genomics Services - Research Branch, Doha, Qatar.
5. Sidra Medicine, Biomedical Informatics – Research Branch, Doha, Qatar.
6. Sidra Medicine, Human Genetics Department, Doha, Qatar.
7. Bioinformatics Core, Weill Cornell Medicine-Qatar, Education City, Doha, Qatar.
8. College of Health and Life Sciences, Hamad Bin Khalifa University, Education City, Doha, Qatar.
9. Qatar Biomedical Research Institute (QBRI), Hamad Bin Khalifa University, Doha, Qatar
10. Molecular Genetics Laboratory, Hamad Medical Corporation, Doha, Qatar.
11. Sidra Medicine, Maternal and Child Health Program, Doha Qatar.
12. College of Health Sciences, Qatar University, Doha, Qatar.
13. Departments of Genetic Medicine, Microbiology and Immunology, Weill Cornell Medicine-Qatar, Doha, Qatar.
14. Sidra Medicine, Clinical Research Center, Doha, Qatar.
15. Center of Genomic and Experimental Medicine, University of Edinburgh, Edinburgh, UK.
16. Biomedical Research Center, Qatar University, Doha, Qatar
17. Department of Genetic Medicine, Weill Cornell Medicine, New York, U.S.A.
18. Department of Molecular Biology and Genetics, Cornell University, New York, U.S.A.

## Acknowledgments

The Qatar Genome Program (QGP) and Qatar Biobank (QBB) are both Research and Development entities within Qatar Foundation for Education, Science and Community Development. We are thankful for everyone who contributed to this endeavor including the QGP and QBB team members, in addition to our partners at Hamad Medical Corporation (HMC), Sidra Medicine and other national stakeholders. We would like to especially thank all participants in this study for their continuous support.

## References Pipelines

QGP: https://qatargenome.org.qa

QBB: https://www.qatarbiobank.org.qa

Sidra Medicine: https://www.sidra.org

dbSNP build 151: http://www.ncbi.nlm.nih.gov/SNP/

HGMD: http://www.hgmd.cf.ac.uk/ac/index.php

ClinVar: https://www.ncbi.nlm.nih.gov/clinvar/

gnomAD: https://gnomad.broadinstitute.org/

GME Variome: http://igm.ucsd.edu/gme/Genomics England:

https://www.genomicsengland.co.uk/

Hail-0.2.13: https://github.com/hail-is/hail/releases/tag/0.2.13

OMIM: https://www.omim.org/

## Data and Code Availability

The informed consent given by the study participants does not cover posting of participant level phenotype and genotype data of Qatar Biobank/Qatar Genome Project in public databases. However, access to QBB/QGP data can be obtained through an established ISO-certified process by submitting a project request at https://www.qatarbiobank.org.qa/research/how-apply which is subject to approval by the QBB IRB committee.

