## Supplemental data for "Qatar Genome: Insights on Genomics from the Middle East" (from another version of this preprint: version 1, 2021-09-23)

**Figure S1. Genomic Quality Score distribution (GQS).** The distribution shows GQS >3.5 across 50 samples from QGP phase I.

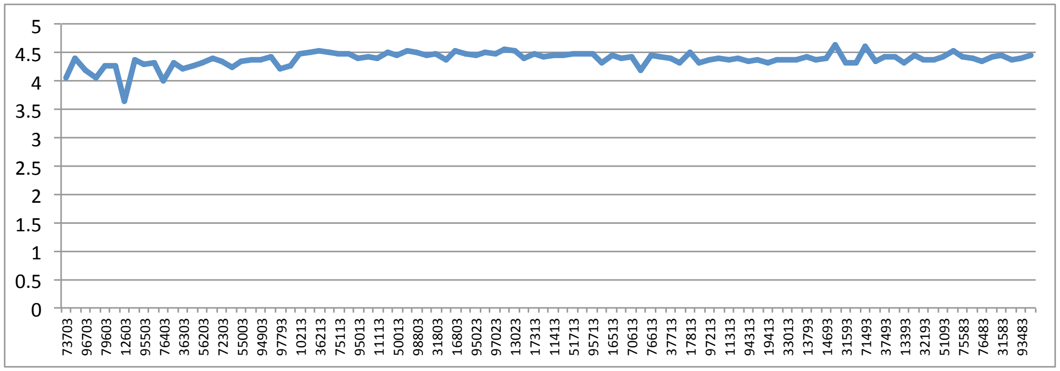


**Figure S2**. **Whole Genome analysis pipeline.** This pipeline follows the GATK 3.4 best practices guidelines. Reads are aligned to hs37d5 reference genome,using bwa.kit. Small variants are called using GATK HaplotypeCaller/ GenotypeGVCFs. Copy number variants are called using Canvas. Structural variants are called using Manta and Delly.

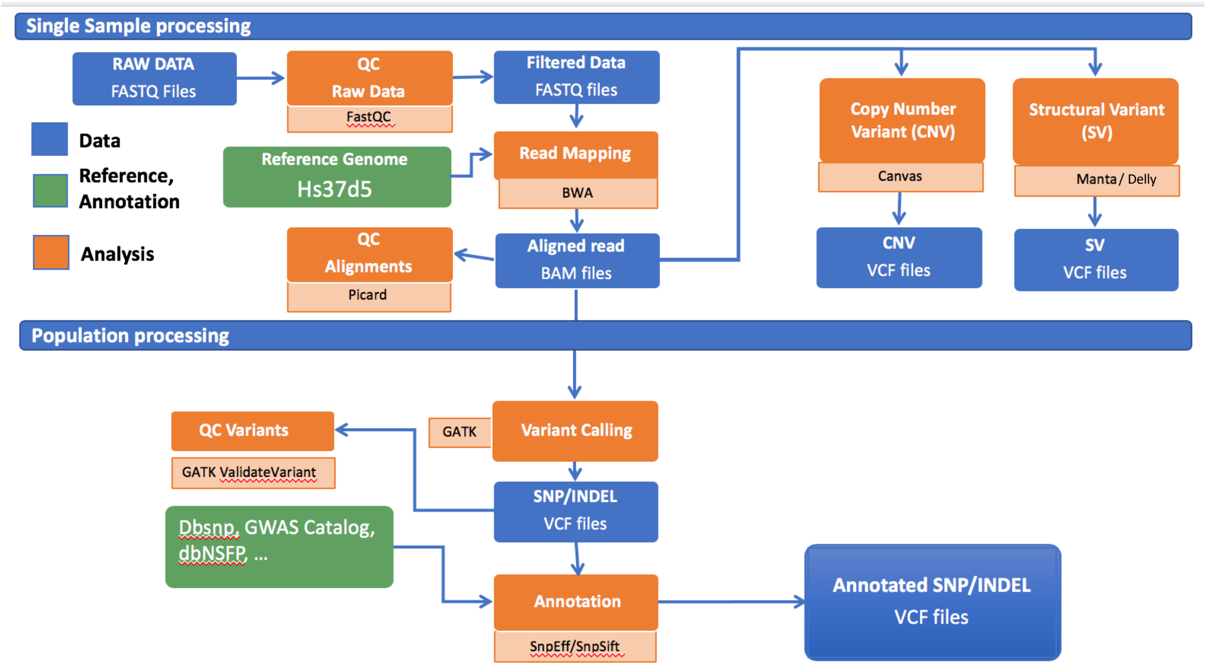

**Figure S3. Variants distribution based on annotation impact and annotation effect.** This annotation impact is obtained by SnpEff/SnpSift tools. High impact has a total of 56,665 variants, Moderate have 587,523, LOW have 1,036,358 and, Modifier have 86,510,692.

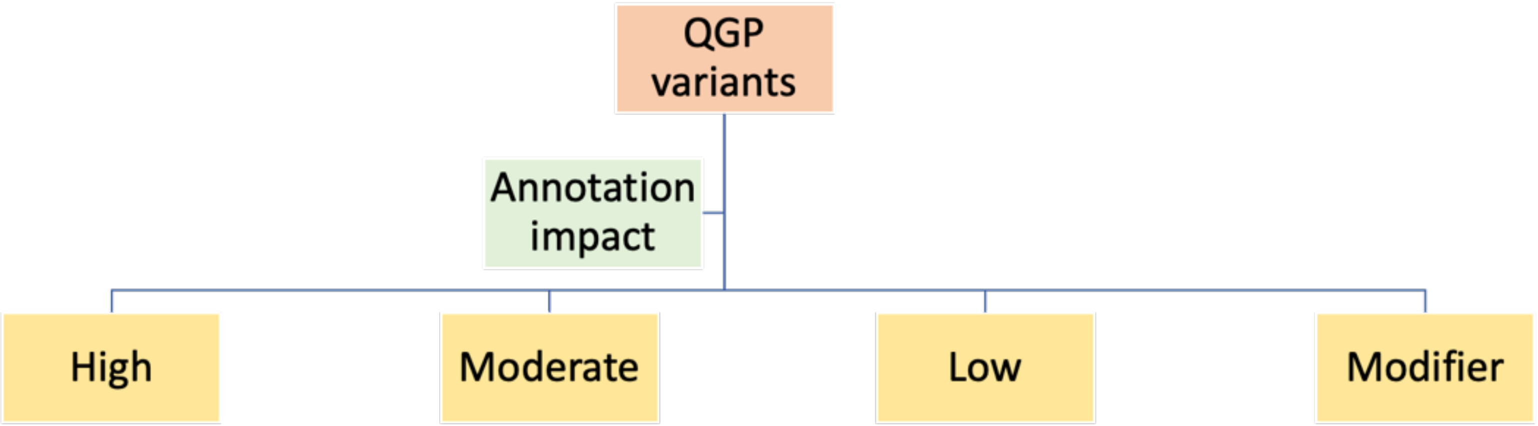


| Annotation effect | No.of variants |
|---|---|
| Exon loss variant | 21 |
| Frameshift variant | 12602 |
| Rare aminoacid variant | 1 |
| Splice acceptor variant | 7219 |
| Splice donor variant | 8695 |
| Start lost | 1570 |
| Stop gained | 10922 |
| Stop lost | 1075 |
| Transcript ablation | 11 |
| Structural interaction variant | 13661 |
| Gene fusion | 223 |
| Protein-protein contact | 434 |
| Bidirectional gene fusion | 231 |

| Annotation effect | No.of variants |
|---|---|
| 5' UTR truncation | 12 |
| Conservative inframe deletion | 2813 |
| Conservative inframe insertion | 1878 |
| Disruptive inframe deletion | 5750 |
| Disruptive inframe insertion | 2510 |
| Missense variant | 484088 |
| Splice region variant | 166 |
| Sequence feature | 90306 |

| Annotation effect | No.of variants |
|---|---|
| 5' UTR premature start codon gain variant | 47419 |
| Initiator codon variant | 111 |
| Splice region variant | 127035 |
| Stop retained variant | 356 |
| Synonymous variant | 285719 |
| TFBS ablation | 2175 |
| TF binding site variants | 37988 |
| Sequence feature | 535555 |

| Annotation effect | No.of variants |
|---|---|
| 3'UTR variant | 950631 |
| 5'UTR variant | 253236 |
| Downstream gene variant | 8704988 |
| Intergenic region | 32662065 |
| Intragenic variant | 7045 |
| Intron variant | 31644914 |
| Non-coding transcript exon variant | 237406 |
| TF binding site variant | 45191 |
| Upstream gene variant | 12005216 |

**Figure S4. Distribution of known and novel SNVs and INDELS of QGP data**. (a) Known and novel SNVs. (b) Known and novel INDELS. 27% of SNV's and 30% of bi-allelic INDELS are novel and not reported in dbSNP build 151 or other population databases (gnomAD, 1000G, GME)

**a)**

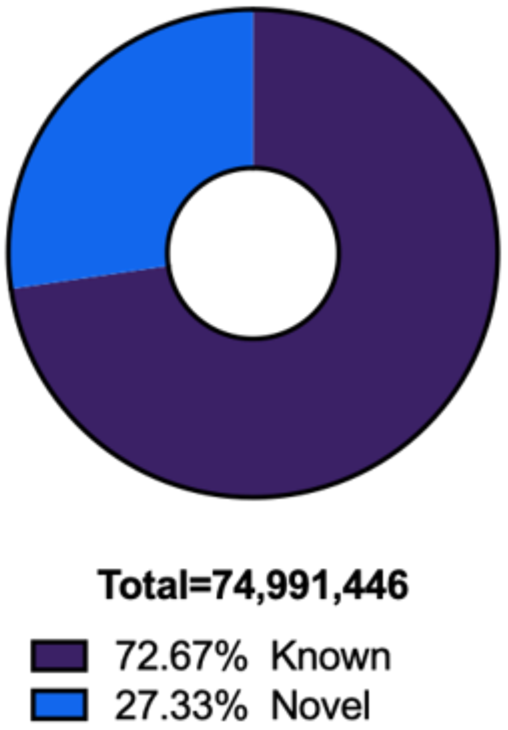


**b)**

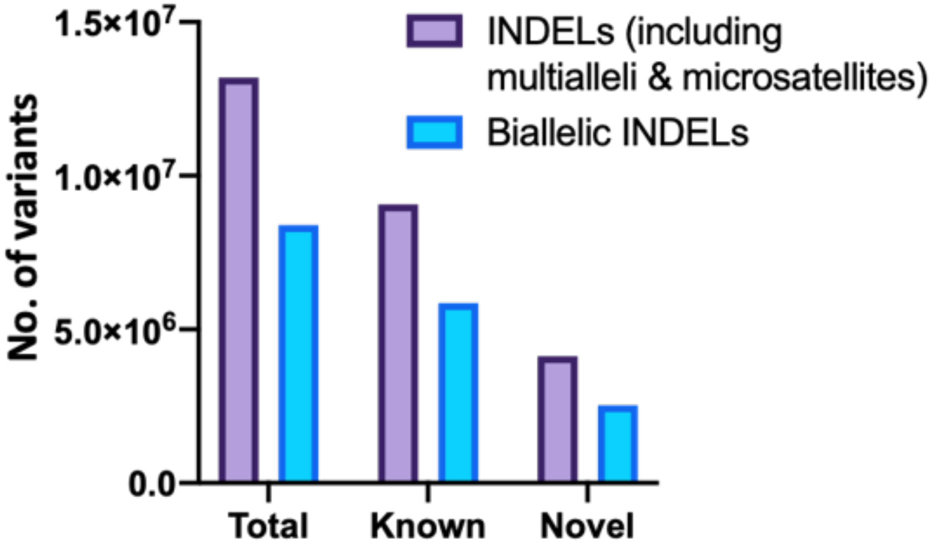

**Figure S5. Comparisons of QGP variants with other population databases.** QGP has an overlap of 55M variants with dbSNP (build151), 46M variants with gnomAD and 244,233 variants with GME database.

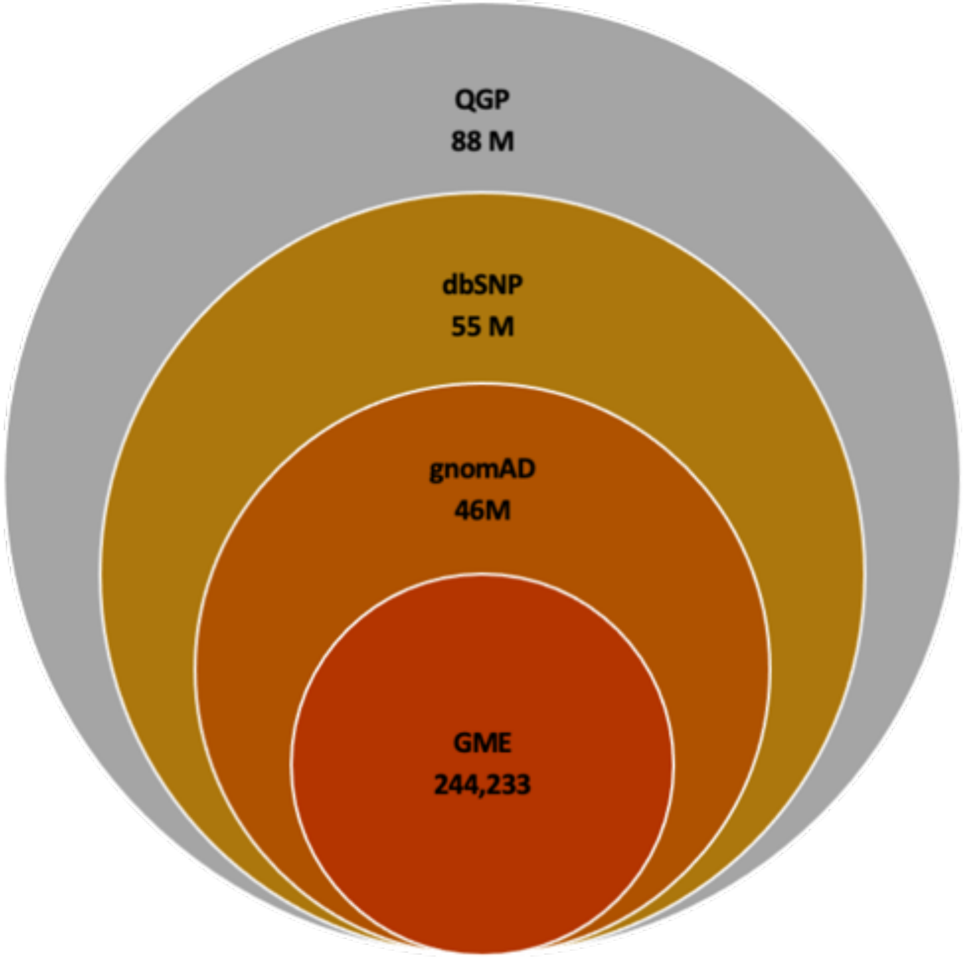

**Figure S6. Allele frequency spectrum of novel SNVs and INDELS.** Novel SNVs are found to be more in the singletons and 2-11 alleles categories, whereas novel INDELs were more common in the MAF categories,1-5%, 5-10% and, >10%. MAF 0.1-1% have a more or less equal number of SNVs (45%) and INDELS (55%).

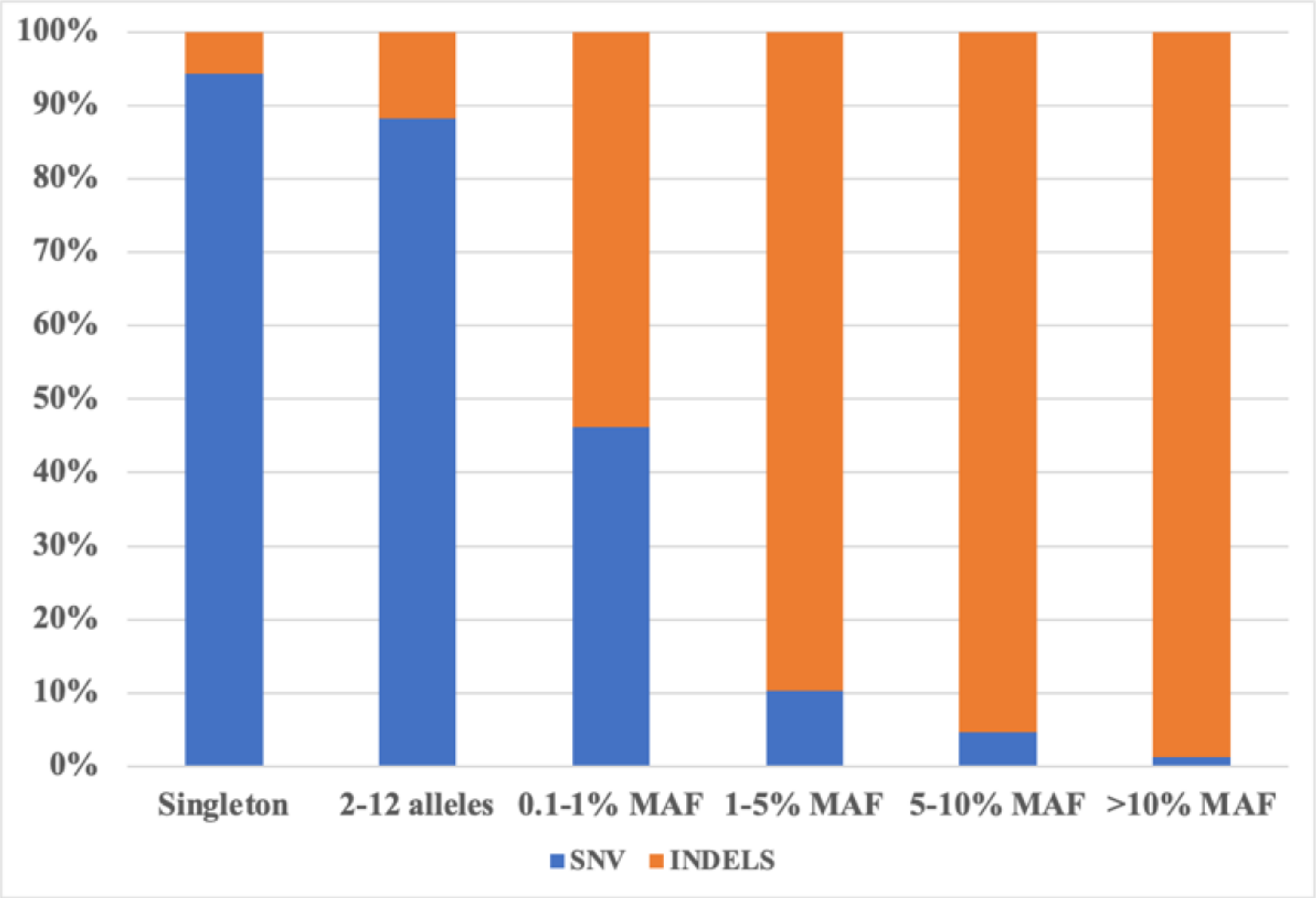

**Figure S7. Variants classification based on HGMD and ClinVar databases.** (a) Functional classes of HGMD variants in QGP data. (b) MAF spectrum of HGMD variants in QGP. (c) Distribution of DM and pathogenic/likely pathogenic variants among QGP individuals. (d) Genes with highest number of DM variants in QGP data. (e) Different classes of ClinVar variants present in QGP data as recommended by ACMG/AMP interpreted for Mendelian disorders.

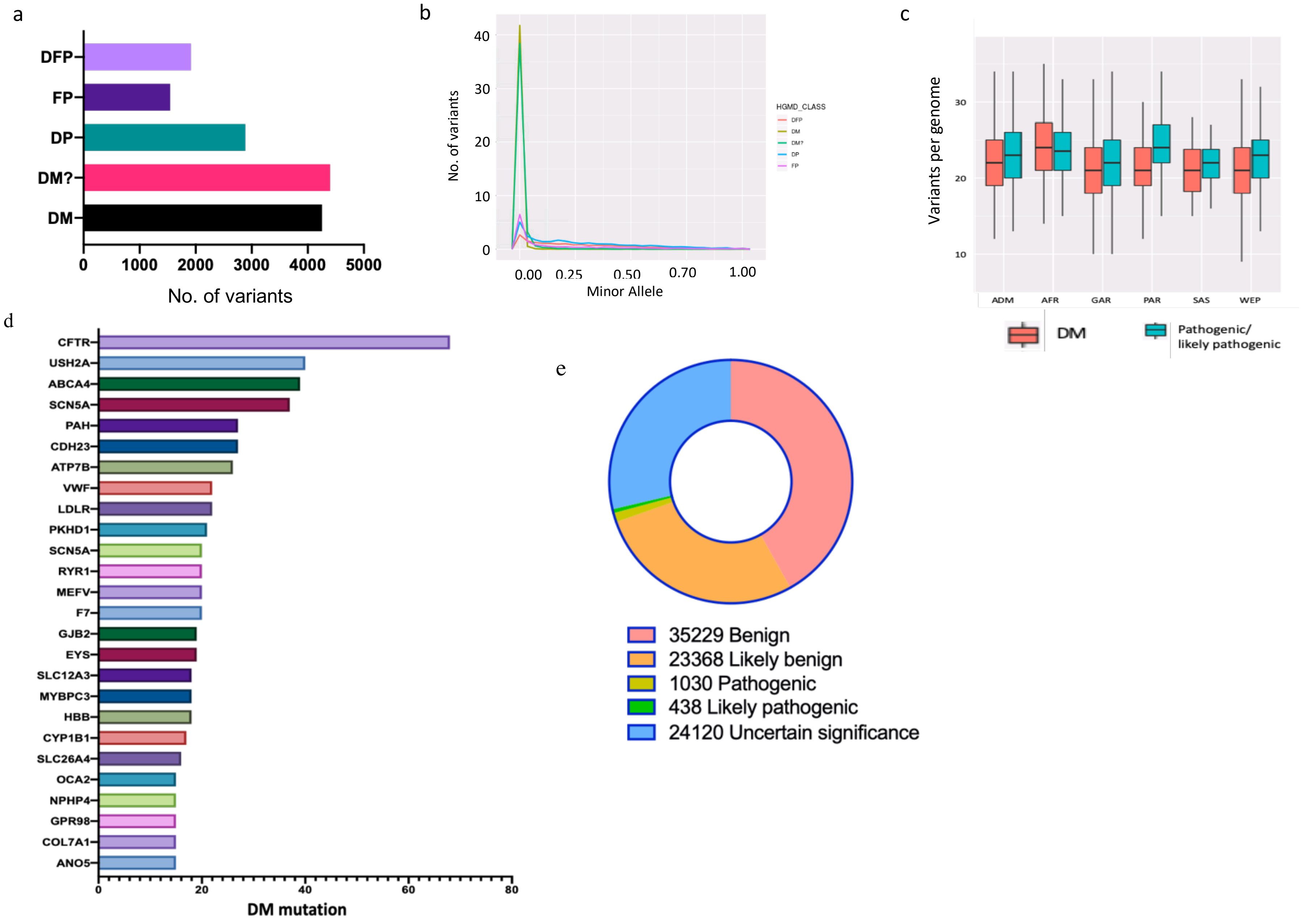

**Figure S8. Population structure of the Qatari population.** a) Average ancestral fractions for QGP and each 1KG population. Results are shown for the k = 8 from the Admixture analysis. b) Admixture bars plot for each QGP individual. Admixed individuals are shown in grey.

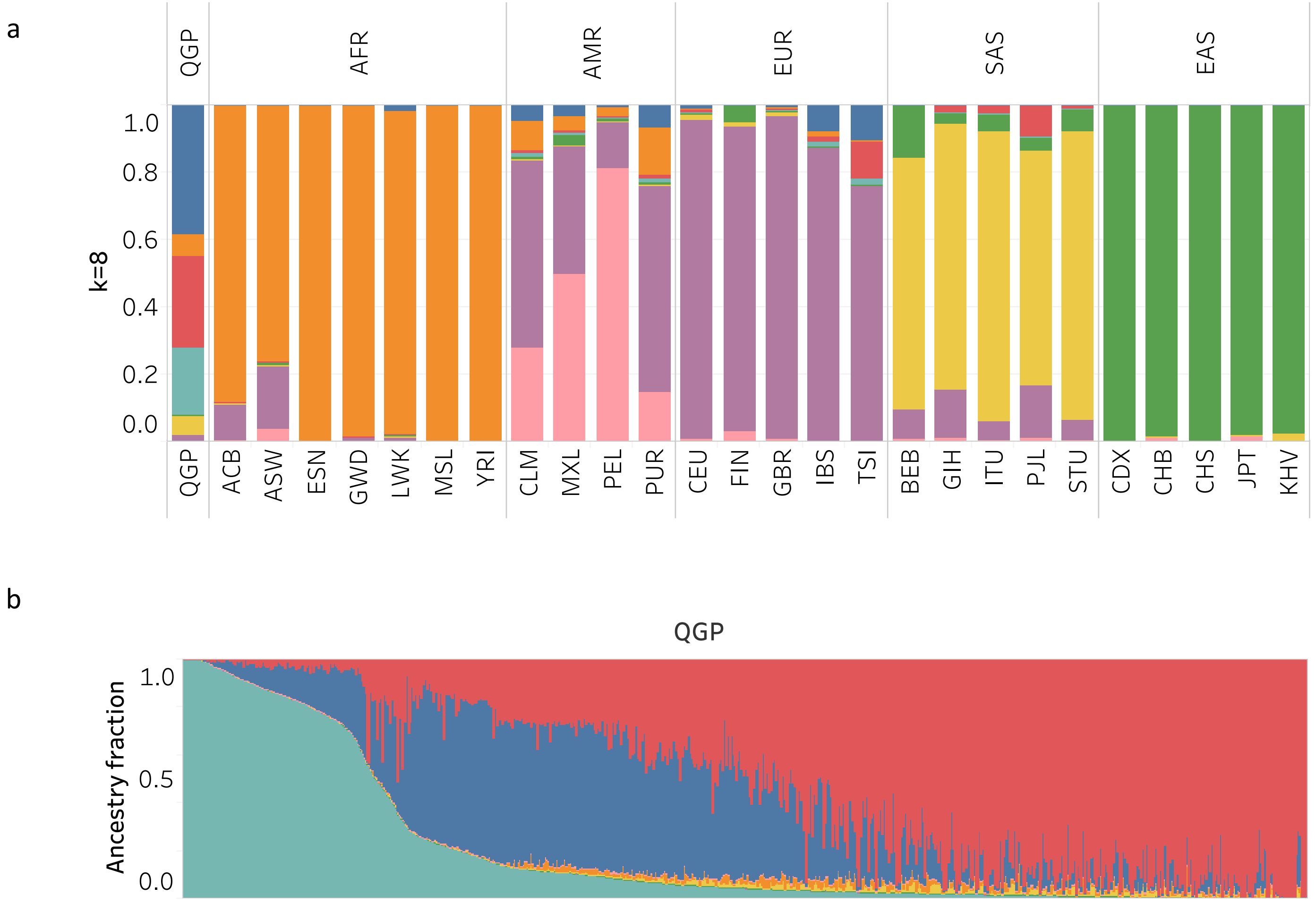

**Figure S9**. **Cluster specific genetic variation.** (a) Novel variants unique to each sub-cluster. (b) Venn diagram of DM variants shared between QGP sub-clusters. (c) Heterozygotes to non-ref homozygotes (het/hom) ratio for the QGP sub-clusters

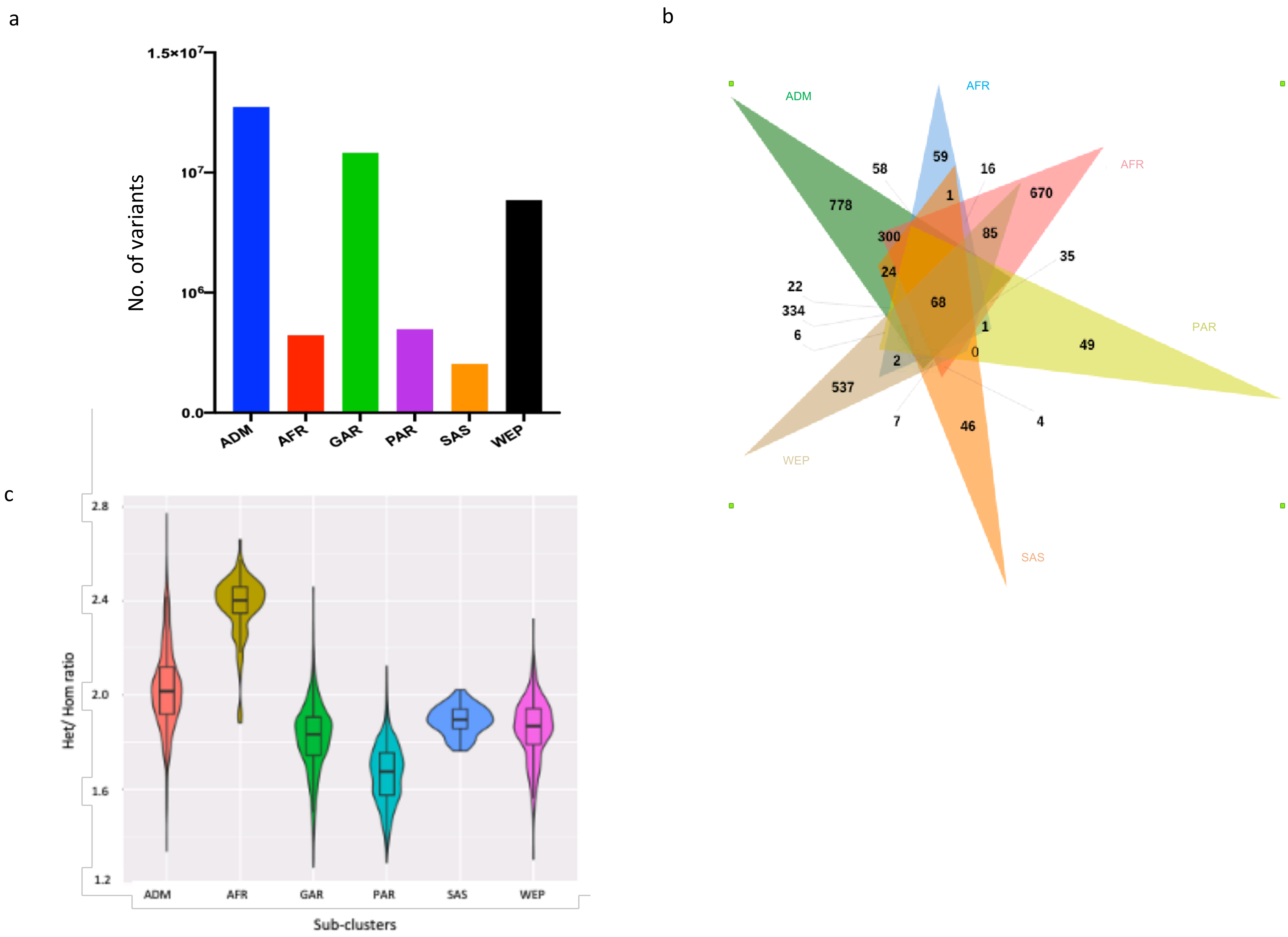

**Figure S10. Association of rs797045038 with Insulin.** The heterozygous individuals (n=87) for rs797045038 show a lower insulin level compared to individuals with a homozygous reference genotype.

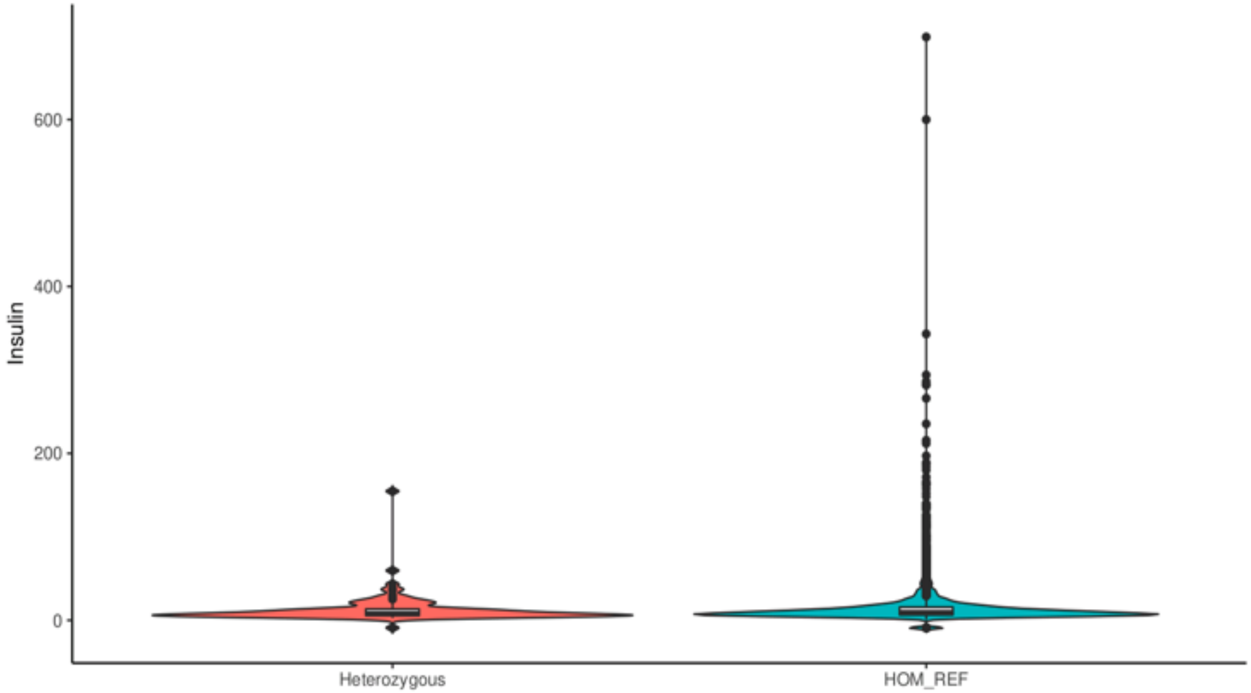

**Table S1**. **QGP variants classification based on annotation impact.**

| | HIGH | MODERATE | LOW | MODIFIER | TOTAL |
|---|---|---|---|---|---|
| Singletons | 19,441 | 206880 | 299340 | 23,338,811 | 23,864,472 |
| <0.1 MAF (2-12 alleles) | 23850 | 251398 | 408411 | 33,377,866 | 34,061,525 |
| 0.1-1 MAF | 8010 | 80003 | 175364 | 15,351,124 | 15,614,501 |
| 1-5 MAF | 2178 | 20060 | 58966 | 5,462,060 | 5,543,264 |
| 5-10 MAF | 768 | 6549 | 21823 | 2,054,239 | 2,083,379 |
| >10 MAF | 2418 | 22633 | 72454 | 6,926,592 | 7,024,097 |
| Total | 56665 | 587523 | 1036358 | 86,510,692 | 88,191,238 |

**Table S2. ClinVar protective variants present in QGP data.**

| CHROM | POS | ID | QGP_MAF | GENE | HGVS_C | HGVS_P | Phenotypes |
|---|---|---|---|---|---|---|---|
| 1 | 67705958 | rs11209026 | 0.051348 | IL23R | c.1142G>A | p.Arg381Gln | Protection against inflammatory bowel disease and psoriasis. |
| 1 | 159174683 | rs2814778 | 0.381925 | DARC | c.-67T>C | . | Protection against *Plasmodium vivax infection.* |
| 1 | 223285200 | rs5744168 | 0.073189 | TLR5 | c.1174C>T | p.Arg392* | Resistance to Systemic Lupus Erythematosus and melioidosis. |
| 2 | 202149589 | rs1045485 | 0.066975 | CASP8 | c.853G>C | p.Asp285His | Protection against breast cancer |
| 3 | 46399208 | rs1799864 | 0.113775 | CCR2 | c.190G>A | p.Val64Ile | Resistance to Human Immunodeficiency Virus (HIV) type 1 |
| 3 | 46411935 | rs1799987 | 0.649992 | CCR5 | c.-2459A>G | . | Resistance to Human Immunodeficiency Virus (HIV) type 1 |
| 3 | 46414573 | rs1800940 | 0.000496 | CCR5 | c.180G>T | p.Arg60Ser | Resistance to Human Immunodeficiency Virus (HIV) type 1 |
| 4 | 100229017 | rs2066702 | 0.012734 | ADH1B | c.1108C>T | p.Arg370Cys | Protection against alcohol dependence |
| 4 | 100260789 | rs698 | 0.383331 | ADH1C | c.1048A>G | p.Ile350Val | Protection against alcohol dependence |
| 4 | 187004074 | rs3775291 | 0.252316 | TLR3 | c.1234C>T | p.Leu412Phe | Resistance to Human Immunodeficiency Virus (HIV) type 1 |
| 5 | 169810796 | rs11739136 | 0.097321 | KCNMB1 | c.193G>A | p.Glu65Lys | Resistance to diastolic hypertension |

| | | | | | | | |
|---|---|---|---|---|---|---|---|
| 6 | 6318795 | rs5985 | 0.115098 | F13A1 | c.103G>T | p.Val35Leu | Protection against myocardial infarction and venous thrombosis |
| 6 | 31910938 | rs547154 | 0.059709 | C2 | n.-512G>T | . | Reduced risk of age-related macular degeneration, 14 |
| 6 | 31914180 | rs641153 | 0.057393 | CFB | c.1601G>A | p.Arg534Gln | Reduced risk of age-related macular degeneration, 14 |
| 7 | 150690079 | rs2070744 | 0.710766 | NOS3 | c.-813C>T | . | Lower risk of developing metabolic syndrome in patients with schizophrenia |
| 8 | 118165323 | rs200185429 | 0.000165 | SLC30A8 | c.412C>T | p.Arg138* | Protection against type 2 diabetic mellitus |
| 9 | 132580901 | rs1801968 | 0.138416 | TOR1A | c.646G>C | p.Asp216His | Protection against dystonia 1 |
| 10 | 43625686 | rs3026785 | 0.059709 | RET | c.*1969T>C | . | Protection against Hirschsprung disease |
| 11 | 116701354 | rs138326449 | 0.000413 | APOC3 | c.109+1G>A | . | Protection against coronary heart disease |
| 11 | 126162843 | rs8177374 | 0.140566 | TIRAP | c.539C>T | . | Protection against invasive pneumococcal disease, bacteremia, malaria and tuberculosis. |
| 13 | 108863609 | rs1805389 | 0.056144 | LIG4 | c.8C>T | p.Ala3Val | Resistance to multiple myeloma |
| 16 | 1129586 | rs121917877 | 8.27E-05 | SSTR5 | c.718C>T | p.Arg240Trp | Resistance to somatostatin analog in |
| 16 | 27356203 | rs1805010 | 0.450885 | IL4R | c.223A>G | p.Ile75Val | Resistance to atophy; and slow progression to acquired immunodeficiency syndrome |
| 16 | 27374180 | rs1805015 | 0.16595 | IL4R | c.1507T>C | p.Ser503Pro | Resistance to atophy |
| 18 | 58038832 | rs52820871 | 0.002729 | MC4R | c.751A>C | p.Ile251Leu | Reduced risk of obesity |
| 9 | 107620867 | rs2230806 | 0.408219 | ABCA1 | c.656G>A | p.Arg219Lys | Reduced risk for premature coronary heart disease in familial hypercholesterolemia |
| 11 | 5248173 | rs33950507 | 0.000496 | HBB | c.79G>A | . | Resistance to malaria |
| 11 | 5248232 | rs334 | 0.00554 | HBB | c.20A>T | p.Glu7Val | Resistance to malaria |
| 11 | 116701353 | rs76353203 | 0.013395 | APOC3 | c.109C>T | p.Arg37* | Reduced risk of coronary heart disease |
| 13 | 108863591 | rs1805388 | 0.100463 | LIG4 | c.26C>T | . | Resistance to multiple myeloma |
| 15 | 99454613 | rs33958176 | 0.000248 | IGF1R | c.1532G>A | p.Arg511Gln | Resistance to insulin-like growth factor I |
| 18 | 58039276 | rs2229616 | 0.010997 | MC4R | c.307G>A | p.Val103Ile | Resistance to obesity |
| X | 138633280 | rs6048 | 0.25831 | F9 | c.580A>G | p.Thr194Ala | Protection against deep venous thrombosis |
| 3 | 46414943 | rs333 | 0.009426 | CCR5 | c.554_585delGTCAGTATCAATTCTGGAAGAATTTCCAGACA | p.Ser185fs | Resistance to Human Immunodeficiency Virus(HIV) type 1 and Hepatitis C virus |
| 2 | 202097531 | rs3834129 | 0.348685 | CASP8 | c.-33678_-33673delAGTAAG | . | Protection against lung cancer |
| 4 | 100504664 | rs1277291635 rs3816873 | 0.386969 | MTTP | c.464T>C | p.Ile155Thr | Protection against metabolic syndrome |

### Table S3: HGMD/ClinVar pathogenic variants with at least 1 homozygous carrier in phase I QGP data

| HGMD disease/phenotype | gene | MIM | variant | dbSNP-id | hom$ | Qatari minor-allele frequency | | | | | Qatar max# | Max subgroup | Global max* |
|---|---|---|---|---|---|---|---|---|---|---|---|---|---|
| | | | | | | PAR | GAR | AFR | SAS | WEP | | | |
| **Desbuquois dysplasia type 2** | XYLT1 | 608124 | c.1588-3C>T; | rs201818754 | 13 | 0.087928 | 0.013631 | 0 | 0 | 0.004009 | 0.087928 | PAR | 0.006623 |
| **Primordial dwarfism** | RBBP8 | 604124 | c.298C>T;p.Arg100Trp | rs373804633 | 10 | 0.067555 | 0.001947 | 0.021739 | 0 | 0.002915 | 0.067555 | PAR | 0.000267 |
| **Autism** | GRIP1 | 604597 | c.160G>A;p.Val54Ile | rs199768740 | 15 | 0.043726 | 0.047815 | 0.005435 | 0 | 0.005102 | 0.047815 | GAR | 0.013245 |
| **Arterial tortuosity syndrome** | SLC2A10 | 606145 | c.243C>G;p.Ser81Arg | rs80358230 | 3 | 0.040875 | 0.001731 | 0 | 0 | 0 | 0.040875 | PAR | 0 |
| **Glaucoma primary open angle/pseudoexfoliation syndrome** | LTBP2 | 602091 | c.1295C>T;p.Pro432Leu | rs137854861 | 1 | 0.009506 | 0.002813 | 0.005435 | 0.039474 | 0.005102 | 0.039474 | SAS | 0.0102 |
| **Homocystinuria** | CBS | 236200 | c.1058C>T;p.Thr353Met | rs398123151 | 2 | 0.033745 | 0.003245 | 0 | 0 | 0 | 0.033745 | PAR | 0.0000359 |
| **Erythrocytosis idiopathic** | SH2B3 | 605093 | c.622G>C;p.Glu208Gln | rs202080221 | 2 | 0.030259 | 0.016681 | 0 | 0 | 0.004389 | 0.030259 | PAR | 0.005032 |
| **Fanconi anemia** | FANCD2 | 613984 | c.2605+1G>A; | rs142365855 | 1 | 0.028992 | 0 | 0.005435 | 0 | 0 | 0.028992 | PAR | 0.0000656 |
| **Griscelli syndrome type 2, Immunodeficiency** | RAB27A | 603868 | c.244C>T;p.Arg82Cys | rs753966933 | 2 | 0.028517 | 0.001515 | 0 | 0 | 0 | 0.028517 | PAR | 0.000183 |
| **2-methylbutyryl-coa dehydrogenase deficiency** | ACADSB | 600301 | c.303+3A>G; | | 2 | 0.025665 | 0.000433 | 0.005435 | 0 | 0.000364 | 0.025665 | PAR | 0.0000325 |
| **Microphthalmia cataract and Iris abnormality** | VSX2 | 142993 | c.599G>C;p.Arg200Pro | rs121912543 | 2 | 0.020437 | 0.001298 | 0 | 0 | 0.000364 | 0.020437 | PAR | 0 |
| **Primary ciliary dyskinesia-13** | DNAAF1 | 613190 | c.1698+1G>A; | rs139519641 | 4 | 0.00903 | 0.017092 | 0 | 0 | 0.001822 | 0.017092 | GAR | 0.002437 |
| **Retinitis pigmentosa** | EYS | 612424 | c.6050G>T;p.Gly2017Val | rs868349465 | 2 | 0.013783 | 0.010169 | 0.01087 | 0 | 0.000364 | 0.013783 | PAR | 0.001025 |
| **Mutilating sensory neuropathy with spastic paraplegia** | CCT5 | 610150 | c.440A>G;p.His147Arg | rs118203986 | 3 | 0.013783 | 0.000649 | 0 | 0.013158 | 0 | 0.013783 | PAR | 0.0043 |
| **Glaucoma primary congenital** | CYP1B1 | 601771 | c.182G>A;p.Gly61Glu | rs28936700 | 2 | 0.010456 | 0.013414 | 0.005435 | 0 | 0.004009 | 0.013414 | GAR | 0.001306 |
| **Intellectual disability, Al-Raqad syndrome** | DCPS | 610534 | c.947C>T;p.Thr316Met | rs137941190 | 1 | 0 | 0.001515 | 0 | 0.013158 | 0.002551 | 0.013158 | SAS | 0.0102 |
| **Congenital adrenal insufficiency** | CYP11A1 | 118485 | c.940G>A;p.Glu314Lys | rs6161 | 1 | 0 | 0.000216 | 0 | 0 | 0.012755 | 0.012755 | WEP | 0.005092 |
| **Polynuropathy, retinal dystrophy, and hearing loss** | ABHD12 | 613599 | c.31C>T;p.Gln11* | rs745990956 | 1 | 0 | 0.001082 | 0.01087 | 0 | 0.00328 | 0.01087 | AFR | 0.0000975 |
| **Dilated cardiomyopathy, muscular dystrophy** | FKTN | 607440 | c.1371_1381dupTATCCAGTTAT;p.Tyr461fs | rs727502847 | 1 | 0.000951 | 0.009736 | 0 | 0 | 0.003644 | 0.009736 | GAR | 0 |
| **Acral peeling skin syndrome 2** | TGM5 | 603805 | c.1335G>C;p.Lys445Asn | rs606231276 | 1 | 0.001426 | 0.008438 | 0.005435 | 0 | 0 | 0.008438 | GAR | 0.000238 |
| **Bamforth-lazarus syndrome** | FOXE1 | 602617 | c.743C>G;p.Ala248Gly | rs538912281 | 1 | 0 | 0.001086 | 0 | 0 | 0.008065 | 0.008065 | WEP | 0.003773 |
| **Hypothyroidism, congenital, nongoitrous, 1** | TSHR | 603372 | c.202C>T;p.Pro68Ser | rs142063461 | 2 | 0.001903 | 0.007799 | 0.005495 | 0 | 0.004383 | 0.007799 | GAR | 0.00183 |
| **Retinitis pigmentosa** | EYS | 612424 | c.2137+1G>A; | rs199740930 | 1 | 0.007605 | 0.007572 | 0 | 0 | 0.000364 | 0.007605 | PAR | 0.009934 |
| **Epidermolysis bullosa dystrophica** | COL7A1 | 120120 | c.4373C>T;p.Pro1458Leu | rs79378857 | 1 | 0 | 0 | 0 | 0 | 0.004373 | 0.004373 | WEP | 0.005102 |
| **Deafness** | OTOF | 603681 | c.5375G>A;p.Arg1792His | rs111033349 | 1 | 0 | 0.001947 | 0 | 0 | 0 | 0.001947 | GAR | 0.000182 |
| **3-methylcrotonyl-coa carboxylase 2 deficiency** | MCCC2 | 609014 | c.538C>T;p.Arg180* | rs780011606 | 1 | 0 | 0.001515 | 0 | 0 | 0 | 0.001515 | GAR | 0.000058 |

$Number of Individual with homozygous genotype
#Maximum minor-allele frequency in local Qatari subpopulation
*Maximum minor-allele frequency in genomAD, ExAC, and 1000 genome variant databases

**Information about the c.436delC founder variant in the *DCAF17* gene in Qatari population**.

| Chr | Locus | Ref | Observed Allele | Variant Type | Gene | dbSNP | Transcript | HGVS.c | HGVS.p | Allele count (heterozygous) | Total allele number | QGP Allele frequency |
|---|---|---|---|---|---|---|---|---|---|---|---|---|
| 2 | 172305304 | TC | T | deletion | DCAF17 | rs797045038 | ENST00000375255.3 | c.436delC | p.Ala147fs | 88 | 12094 | 0.00727 |

**Testing Method:**
Samples are tested using bi-directional targeted Sanger sequencing of the exon encompassing the c.436delC in the DCAF17 gene. Mutation nomenclature is according to Human Genome Variation Society (HGVS) convention with numbering starting at the A of the ATG initiation site. The DCAF17 (NM_025000.3) is used as references.

**References pipelines**


1. http://maq.sourceforge.net/fastq.shtml
2. https://www.bioinformatics.babraham.ac.uk/projects/fastqc/
3. ftp://ftp.1000genomes.ebi.ac.uk/vol1/ftp/technical/reference/phase2_reference_assembly_sequence/hs37d5.slides.pdf


ftp://ftp.1000genomes.ebi.ac.uk/vol1/ftp/technical/reference/phase2_reference_assembly_sequence/hs37d5.slides.pdf


4. https://github.com/lh3/bwa/tree/master/bwakit
5. http://samtools.github.io/hts-specs/
6. https://software.broadinstitute.org/gatk/documentation/tooldocs/4.0.0.0/picard_analysis_CollectWgsMetrics.php
7. https://software.broadinstitute.org/gatk/documentation/article?id=3238
8. https://github.com/Intel-HLS/GenomicsDB
9. http://snpeff.sourceforge.net/
10. https://sites.google.com/site/jpopgen/dbNSFP
11. https://www.genome.gov/gwastudies/
12. http://software.broadinstitute.org/gsea/msigdb/index.jsp
13. https://github.com/Illumina/canvas
14. https://github.com/Illumina/manta
15. https://github.com/dellytools/delly
16. https://genome.sph.umich.edu/wiki/Vt
